# Host-related concordance of TAC/SARIFA in colorectal double and triple carcinomas suggests patient-specific metabolic reprogramming

**DOI:** 10.64898/2026.07.12.26357852

**Authors:** Francisco J. Farfán López, Armin Wiegering, Bruno Märkl, Johanna Waidhauser, Markus Krebs, Bianca Grosser, Nic G. Reitsam, Andreas Probst, Matthias Schrempf, Gerhard Schenkirsch, Andreas Rosenwald, Florian Kurz

## Abstract

**Introduction:** TAC/SARIFA has been introduced as a new robust and easy to evaluate biomarker in several cancer entities including colorectal cancer. It is defined by a direct contact of at least five tumour cells with one adipocyte and is believed to indicate metabolic reprogramming associated with adverse outcome. However, the mechanism that leads to TAC/SARIFA-positivity is unclear, yet. To investigate whether there is an individual’s component we conducted a study on double and triple cancer establishing a within-patient-design.

**Methods:** We retrospectively analysed a total number of 135 cases with 276 colorectal cancers from two academic medical centres. The TAC/SARIFA status was evaluated as well as the basic histopathological factors. The median follow-up time was 120 months.

**Results:** Cases with any TAC/SARIFA-positive tumours showed a significant reduced overall survival (62 v s. 88 months; p = 0 .011). Analysing the entire cohort the rates of concordant and discordant cases followed a random distribution. However, restriction to synchronous pT3/4-cases revealed a significant (p = 0.016) deviation from a random distribution.

**Conclusion:** This study reveals significant concordance of TAC/SARIFA status in synchronous locally advanced colorectal double/triple carcinomas, supporting the concept that tumour–adipocyte interaction reflects a host-related microenvironmental condition linked to metabolic reprogramming rather than a purely tumour-intrinsic event.

## 1 Introduction

Colorectal cancer (CRC) is a heterogeneous tumour entity with a poor prognosis in advanced stages, and despite the identification of specific, partly environmental risk factors, improved preventive measures, and therapeutic strategies, overall patient survival is limited. It has the second-highest mortality rate after lung cancer and, with an incidence of almost two million new cases in 2022, is the third most common cancer worldwide (1). The prognosis estimation and the therapy stratification still rely on the tumour stage based on the criteria of the system according to the UICC/AJCC (2, 3). For stages I to III, only a few additional biomarkers gained relevance and acceptance in daily routine. Due to its predictive values regarding checkpoint inhibitor therapy, evaluating the microsatellite (MSS vs. MSI) or mismatch repair status (pMMR vs. dMMR) (4) is essential not only to detect cases of hereditary non-polyposis colorectal cancer. With the publication of the results of the International Tumour Budding Consensus Conference in 2016 (3), tumour budding (TB) also became part of routine reporting in CRC. Recently, a large study confirmed the prognostic value of TB quantitatively (5) and it is also prognostic in early lesions (5, 6). Despite its widely accepted value for the management of CRC, the method is hampered by a certain interobserver variability, which is not improved by additional cytokeratin immunohistochemistry or deep learning algorithms (7–10). Recently, we introduced TAC/SARIFA (tumour-adipocyte contact / stroma areactive invasion front areas; formerly: SARIFA) as a new histomorphological biomarker in several cancer entities. It is defined by direct contact of at least five tumour cells with at least one adipocyte and reflects metabolic reprogramming of the tumour cells towards a lipid-driven cancer progression. In gastric and colorectal cancers, the biomarker has been investigated most intensively with more than 8.000 evaluated cases, including three reports from other independent groups confirming its strong prognostic power (4–6) (11). TAC/SARIFA is easy and fast to evaluate without the need for special stainings. Moreover, the interobserver variability is very low. Very recently, a consensus meeting was held regarding SARIFA, confirming the definition of SARIFA and its prognostic value, also in the view of the expert panel. It was also recommended that the nomenclature be amended, as SARIFA does not accurately reflect the key feature of the tumour-adipocyte interaction. Tumour-adipocyte contact (TAC) should be incorporated, and therefore, ‘TAC/SARIFA’ should be used as the future designation. For this reason, we will adopt this terminology in the following sections of this article (12). Because metabolic reprogramming is the supposed main mechanism, targeting lipid metabolism is a potential therapeutic option in the future (13). As far as is known, TAC/SARIFA is not driven by genetics (14). Obesity has been suspected to be related to TAC/SARIFA. This could not be confirmed in retrospective studies enrolling cancer cases (15, 16). However, the analysis of the large prospective population-based Netherlands Cohort Study on diet and cancer (NLCS) revealed a higher risk for the development of a SARIFA-positive colorectal cancer for individuals with an increased baseline bodyweight. Next to it, only increased protease activity and impaired immune response could be identified as recurring conditions in TAC/SARIFA-positive tumours (17–20). This fact leads us to the hypothesis that certain features of the affected individual host organisms lead to a lack of stromal reaction and direct interaction between tumour cells and adipocytes. To elucidate the individual’s component, we have chosen a split body design to minimise confounders that are not related to the individual patient. We analysed double and triple colorectal cancers because if the thesis holds true that TAC/SARIFA is triggered by the host and its tumour microenvironment (TME), the TAC/SARIFA-status of the tumours within an individual patient should be identical.

## 2 Materials and Methods

### 2.1 Cohort

The archive files of two academic pathology departments (UK Augsburg and Würzburg) were retrospectively screened for synchronous or metachronous double or triple colorectal cancers diagnosed between 2000 and 2024. The only exclusion criteria were a complete response after neoadjuvant therapy or the absence of a surgical specimen. Especially the early stages (pT1) were included, as were hereditary cancers and cases of inflammatory bowel disease. The follow-up data were provided by the Augsburg tumour registry and the Bavarian tumour registry run by the Landesamt für Gesundheit und Lebensmittelsicherheit. Ethical approval was obtained from the ethics committees of the LMU Munich (reference project number: 22-0120).

### 2.2 Definitions

For comparisons, we defined a first −, a second, and a third tumour, respectively. Regarding that, metachronous cases were classified according to their temporal occurrence. In synchronous cases tumours were classified according to their pT-stage under a simplifying assumption – which does not necessarily reflect the truth - that larger tumours occurred earlier than smaller ones. In cases of equal pT-stage, the tumour with a shorter distance to the Bauhin valve was arbitrarily defined as the “first” tumour. In metachronous cases, both tumours were classified as “metachronous”, which does not correspond to clinical usage, where only the tumour that developed later is referred to as ‘metachronous’.

### 2.3 Histological re-evaluation

All identified cases were histologically re-evaluated by FK and FF, and all available H.E.-stained tumour slides were screened. TAC/SARIFA has been classified according to the original definition of at least 5 tumour cells or one tumour gland with direct contact to at least one adipocyte without intervening stroma or immune cells (21) (Figure 1). Regarding the TAC/SARIFA-status, the Würzburg-collective has been evaluated by four experienced pathologists (AR, FK, BM, and FF), establishing a consensus diagnosis on a multi-head microscope. The Augsburg-collective has been re-evaluated by FF. In difficult cases, a consensus diagnosis has been established together with BM. Regarding the additional histological features (pT-, L-,V-, Pn-stage, Grading, tumour budding) all cases were re-evaluated by FF.

**Figure 1.**
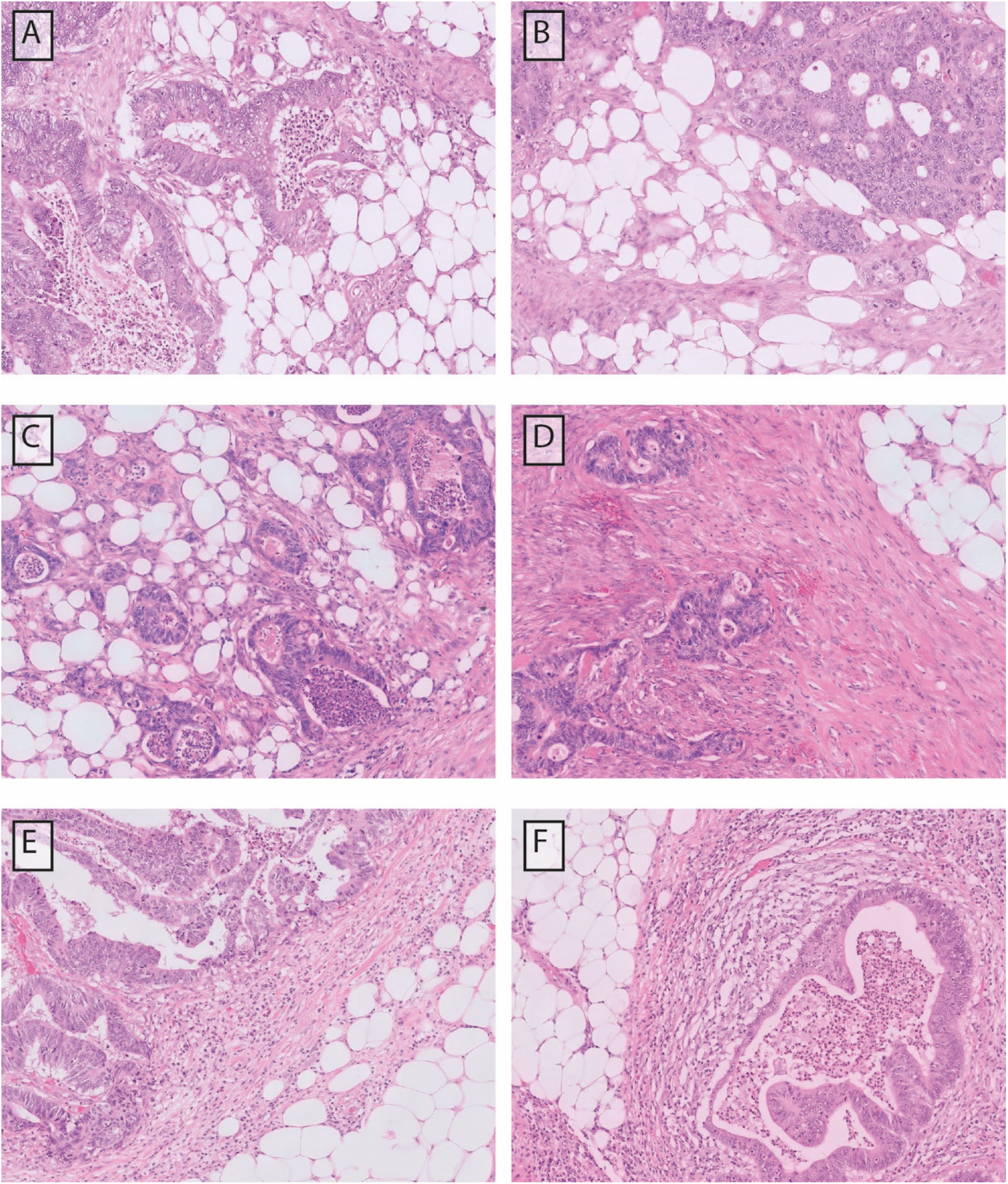
Histopathological Images (HE, Magnification 10x); A and B show concordant TAC/SARIFA-positive tumours. C and D represent a discordant case with TAC/SARIFA-positive tumour in C and a negative tumour in D. E and F are an example of two concordant TAC/SARIFA-negative tumours.

### 2.4 Immunohistochemical evaluation and molecular profiling

In cases where the mismatch-repair (MMR)-status was missing and formalin-fixed and paraffin-embedded (FFPE) material was available, immunohistochemical analyses were performed using antibodies against MLH1 (Clone: M1, Roche, Basel, Switzerland), PMS2 (Clone: EP51, Agilent Technologies, Santa Clara, CA, USA), MSH2 (Clone: G219-1129, Roche, Basel, Switzerland) and MSH6 (Clone: EP49, Leica Biosystems, Newcastle, UK). The reactions were developed using the OptiView DAB IHC Detection Kit on a Roche Ventana benchmark ultra platform (Ventana Medical Systems, Inc., Tucson, AZ, USA; Roche Diagnostics). Molecular data have been obtained from the available reports. No additional molecular analyses have been performed.

### 2.5 Statistics

Continuous variables were compared using the t-test or Wilcoxon rank sum test, depending on the distribution. Tabulated data were compared using either the Chi-square or Fisher’s exact test, depending on the group sizes. Overall survival analyses were performed using the Log-rank-test and Kaplan-Meier-Curves. A multivariate Cox-regression analysis was performed to identify independent parameters. Survival analyses were administratively censored at 120 months. The follow-up time was calculated from the time of the initial diagnosis to the last available time point or death, using the method of Schemper and Smith (22). Expected TAC/SARIFA concordance rates were calculated under a random independence model. The deviation between the observed and expected distributions was assessed using an exact Pearson-type goodness-of-fit test that preserves the observed numbers of double and triple carcinoma cases.

Statistical analyses were performed using SigmaPlot version 16.0.0.28 (Systat Software, Inc., San Jose, CA, USA) and R version 4.3.3 (R Foundation for Statistical Computing, Vienna, Austria). The exact goodness-of-fit test for TAC/SARIFA concordance was implemented using custom R code based on base R and the stats package version 4.3.3. A p-value < 0.05 was considered significant.

## 3 Results

### 3.1 Cohort

A total number of 135 cases with 276 tumours (EntireCohort) were included between 2000 and 2024. The clinico-pathological data are summarized in Table 1. A sub-cohort was extracted, restricted to synchronous double/triple pT3/4 cases (SynchAdvCohort), comprising 31 individuals with a total of 64 tumours (Table 2). The MetaAdvCohort includes 26 patients with 52 metachronous tumours. The Suppl-Table 1 provides an overview of the entire cohort on a tumour basis.

**Table 1.**

| Entire cohort |  |  |  |  |  |
| --- | --- | --- | --- | --- | --- |
|  | All | SARIFA pos<br>concordant | SARIFA pos<br>discordant | SARIFA neg<br>concordant | P-Value |
|  | **n = 135**<br>**n = 276** | **n = 17**<br>**n = 34** | **n = 52**<br>**n = 106** | **n = 66**<br>**n = 136** |  |
| Collect Würzburg | 36 | 5 | 14 | 17 |  |
| Collect Augsburg | 99 | 12 | 38 | 49 |  |
| Synchronous | 91 | 13 | 37 | 41 | 0.405 |
| Metachronous | 44 | 4 | 15 | 25 |  |
| Mean Age (± SD) | 69 ± 12 | 65 ± 9 | 68 ± 15 | 68 ± 12 | 0.873 |
| Sex (M : F) | 1.9 : 1 | 1.4 : 1 | 1.2 : 1 | 2.9 : 1 | 0.090 |
| Double Cancer | 129 | 17 | 50 | 62 | n.a. |
| Triple Cancer | 6 | 0 | 2 | 4 |  |
| Right Colon | 121 | 17 | 46 | 58 | 0.613 |
| Left Colon Rectum | 83 | 10 | 34 | 39 |  |
| Total Colon/Rectum | 58 | 5 | 19 | 34 |  |
| Unknown Location | 1 | 0 | 1 | 0 |  |
| pT1* | 13 | 2 | 6 | 5 |  |
|  | 44 | 1 | 20 | 23 | < 0.001 |
| pT2* | 55 | 1 | 14 | 40 |  |
| pT3* | 140 | 22 | 56 | 62 |  |
| pT4* | 37 | 10 | 16 | 11 |  |
| pN0 | 160 | 8 | 62 | 90 | < 0.001 |
| pN+ | 109 | 26 | 42 | 41 |  |
| low grade | 213 | 22 | 81 | 110 | 0.072 |
| high grade | 57 | 12 | 22 | 23 |  |
| Tumor Budding 1 | 207 | 22 | 70 | 115 | 0.001 |
| Tumor Budding 2/3 | 69 | 12 | 36 | 20 |  |
| MSS / pMMR | 165 | 26 | 67 | 72 | 0.037 |
| MSI / dMMR | 38 | 0 | 17 | 21 |  |
| MSI / dMMR - BRAF WT | 16 | 0 | 9 | 7 | n.a. |
| MSI / dMMR - BRAF mut | 8 | 0 | 3 | 5 |  |
| MS / MMR Discordance | 6 | 0 | 1 | 5 | n.a. |
| Neo-Adjuvant Therapy | 21 | 2 | 4 | 15 |  |

Items in orange text font refer to cases and in blue font to tumours. SD = 1 standard deviation, M = male, F = female, MSS = microsatellite stable, MSI = microsatellite instable, pMMR = proficient mismatch repair, dMMR = deficient mismatch repair, \* T-stage refers to the first tumour.

**Table 2.**

| Synchronous tumours pT3/4 |  |  |  |  |  |
| --- | --- | --- | --- | --- | --- |
|  | All | SARIFA pos<br>concordant | SARIFA pos<br>discordant | SARIFA neg<br>concordant | P-Value |
|  | n = 31<br>n = 64 | n = 12<br>n = 24 | n = 8<br>n = 16 | n = 11<br>n = 24 |  |
| Collect Würzburg Collect | 5 | 2 | 0 | 3 |  |
| Augsburg | 26 | 10 | 8 | 8 |  |
| Mean Age (± SD) | 72 ± 10 | 70 ± 10 | 80 ± 9 | 71 ± 7 | **0.039** |
| Sex (M : F) | 2.1 : 1 | 1.4 : 1 | 1.6 : 1 | 4.5 : 1 | n.a. |
| Double Cancer | 29 | 12 | 8 | 9 | n.a. |
| Triple Cancer | 2 | 0 | 0 | 2 |  |
| Right Colon | 34 | 14 | 8 | 12 | 0.981 |
| Left Colon | 15 | 5 | 4 | 6 |  |
| Rectum | 15 | 5 | 4 | 6 |  |
| Total Colon/Rectum | 0 | 0 | 0 | 0 |  |
| Unknown Location | 0 | 0 | 0 | 0 |  |
| pT3* | 17 | 8 | 3 | 6 | 0.578 |
| pT4* | 47 | 16 | 13 | 18 |  |
| pN0 | 23 | 4 | 6 | 13 | **0.025** |
| pN+ | 41 | 20 | 10 | 11 |  |
| low grade | 44 | 14 | 14 | 16 | 0.134 |
| high grade | 18 | 10 | 2 | 6 |  |
| Tumor Budding 1 | 46 | 15 | 11 | 20 | 0.262 |
| Tumor Budding 2/3 | 18 | 9 | 5 | 4 |  |
| MSS / pMMR | 47 | 20 | 14 | 13 | **0.002** |
| MSI / dMMR | 11 | 0 | 2 | 9 |  |
| MSI / dMMR - BRAF WT | 3 | 0 | 0 | 3 | n.a. |
| MSI / dMMR - BRAF mut | 6 | 0 | 2 | 4 |  |
| MS / MMR Discordance | 1 | 0 | 0 | 1 | n.a. |
| Neo-Adjuvant Therapy | 4 | 1 | 0 | 3 |  |

Items in orange text font refer to cases and in blue font to tumours. SD = 1 standard deviation, M = male, F = female, MSS = microsatellite stable, MSI = microsatellite instable, pMMR = proficient mismatch repair, dMMR = deficient mismatch repair, \* T-stage refers to the first tumour.

### 3.2 Sequential data

The pT-stage distribution of first and second tumours is given in Figure 2. Due to its definition the number of pT3/4-stages in synchronous first tumours is increased 77 vs. 31 while the distribution is balanced in metachronous tumours 34 vs 32.

**Figure 2.**
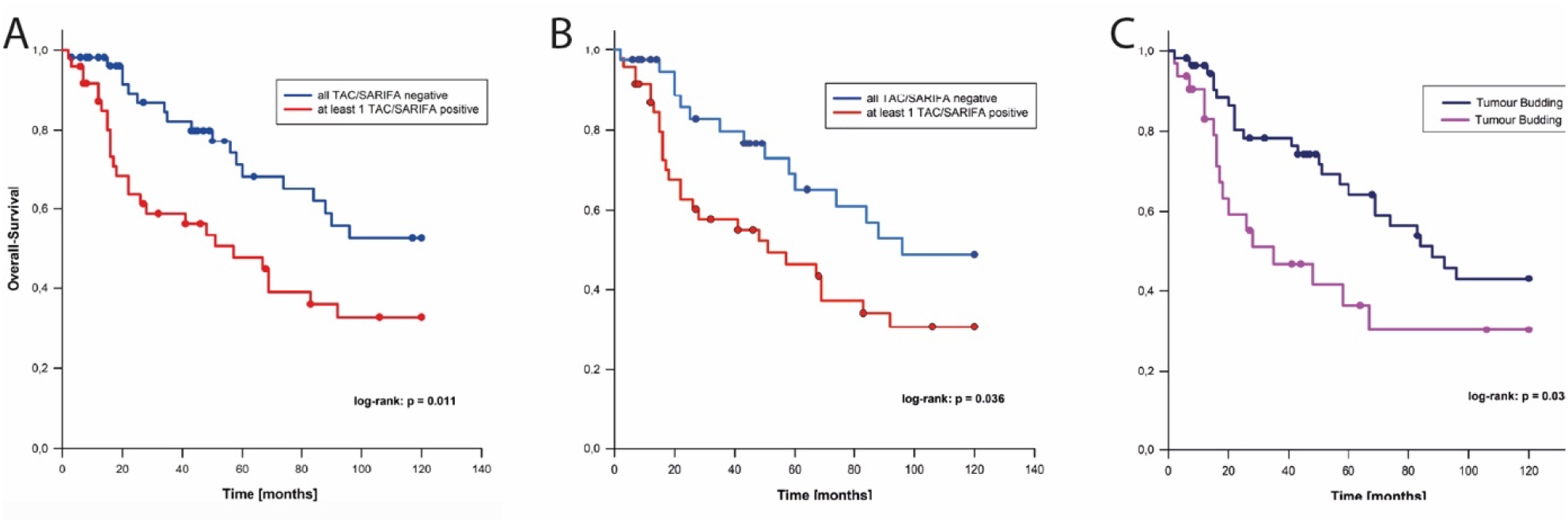
A Overall Survival TAC/SARIFA stratified in the entire cohort; B Overall Survival

### 3.3 Survival

The median follow-up time was 120 months for all analyses regarding TAC/SARIFA and Budding. TAC/SARIFA-positivity was significantly associated with adverse overall survival in the entire cohort (n = 103; mean survival time: 62 vs. 88 months; p = 0.011) (Figure 2A) and a sub-cohort restricted to pT3/4 cases (n = 87; mean survival time: 61 vs. 84 months; p = 0.036) (Figure 2 B). Two-tiered tumour budding showed an almost similar prognostic performance (n = 87; mean survival time: 55 vs. 80 months; p = 0.034) (Figure 2C). In multivariable Cox proportional hazards analysis, high tumour budding remained an independent prognostic factor (HR 2.31, 95 % CI 1.21–4.42, p = 0.012). TAC/SARIFA positivity showed a strong trend towards worse outcome (HR 1.90, 95 % CI 0.99– 3.67, p = 0.054). pT and pN status were not independently associated with outcome.

TAC/SARIFA stratified in pT3/4 cases; C Overall Survival two-tiered tumour budding stratified

### 3.4 TAC/SARIFA-distribution: Entire Cohort

The overall TAC/SARIFA-positivity rate in this cohort of 135 cases was 31%. Under the assumption of a fully random distribution regarding the TAC/SARIFA status, the expected rates for concordant positive, discordant positive/negative and concordant negative cases would be 13 (10%; 95respectively. The rates in the analysed cohort were 17 (13%), 52 (39%) and 66 (49%), and therefore entirely in the expected range of a random distribution (p = 0.306)(Figure 3A). In this cohort, concordant or discordant SARFIA-positivity was associated with higher pT-stage, LN-positivity, higher Tumour Budding grade and mismatch-repair-proficiency (pMMR) / microsatellite stability (MSS) (Table 1).

**Figure 3.**
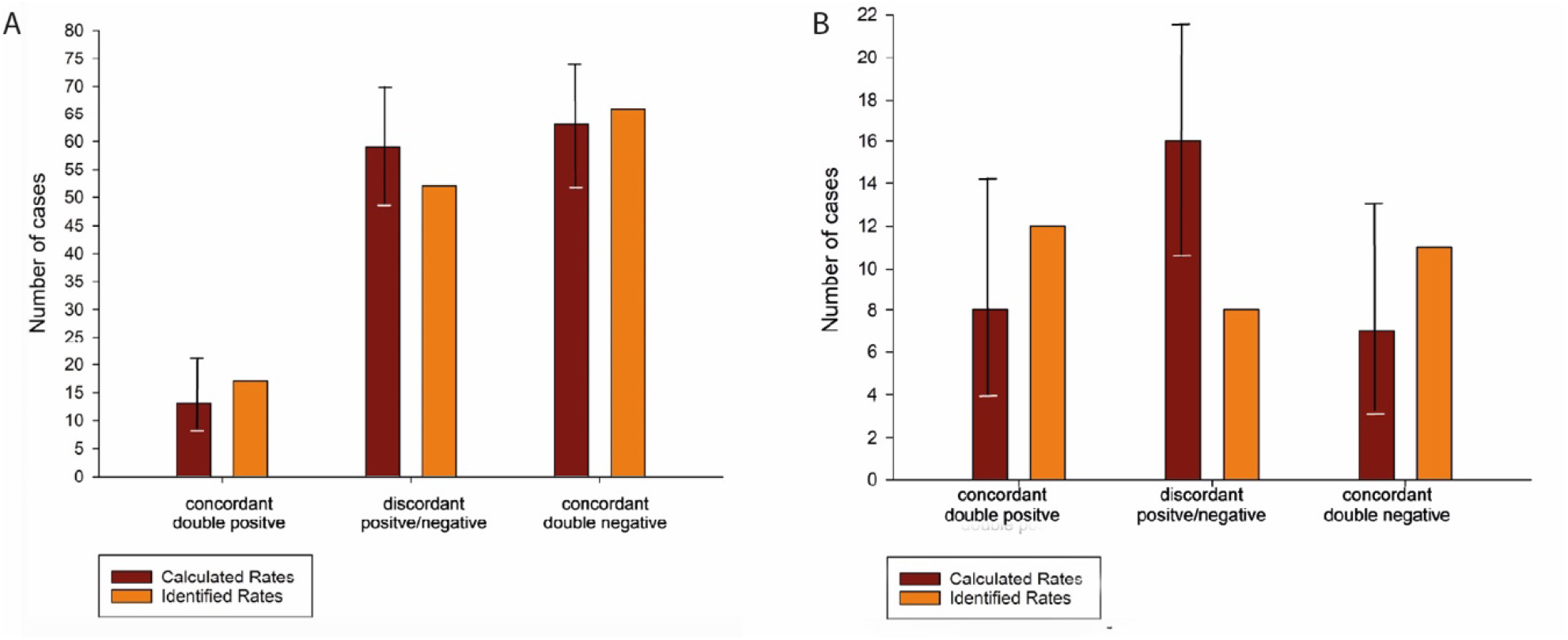
Calculated and identified concordance rates A in Entire Cohort and B in SynchAdvCohort

### 3.5 TAC/SARIFA-distribution: SynchAdvCohort

To exclude the effect of pT stage and fully reduce confounding factors, the SynchAdvCohort has been constructed to include exclusively synchronously developed pT3/4 tumours. This cohort comprises 31 cases, with a TAC/SARIFA-positivity rate of 51%. Under a fully random distribution of TAC/SARIFA status in this cohort, the expected rates for concordant positive, discordant positive/negative, and concordant negative cases would be 8 (26%; 95and 7 (24%; 95cohort were 12 (39%), 8 (26%), and 11 (35%), respectively, indicating an excess of concordant cases and a reduction of discordant cases compared with the random null distribution (p = 0.016). In this cohort, concordance of SARIFA-positivity was associated with higher LN-positivity (p = 0.025) and mismatch-repair proficiency (pMMR)/microsatellite stability (MSS) (p = 0.002) (Table 2). Patients with discordant TAC/SARIFA status were significantly older than those with concordant status (80 years ± 9 vs. 70 years ± 10 and 71 years ± 7, p = 0.039).

### 3.6 TAC/SARIFA-distribution: MetaAdvCohort

Of the 26 metachronous cases, 25 fulfilled the criteria for locally advanced pT3/4 disease and were included in the MetaAdvCohort analysis, which was built to investigate whether discordant cases follow a preferential temporal sequence. This is not the case in the total of 12 discordant cases, with five cases starting with a TAC/SARIFA-negative tumour. The TAC/SARIFA distribution of this cohort with 9 (36%) concordant negative, 12 (48%) discordant and 4 (16%) concordant positive cases is identical with the calculated random distribution: 8 (95

## 4 Discussion

The central finding of the present study is the non-random concordance of TAC/SARIFA status in synchronous locally advanced colorectal carcinomas occurring within the same individual. While the distribution of TAC/SARIFA-positive and TAC/SARIFA-negative tumours across the entire cohort was consistent with a random occurrence, restriction to synchronous pT3/pT4 carcinomas revealed a significant excess of concordant cases. This observation supports the hypothesis that tumour-adipocyte contact is not exclusively determined by tumour-intrinsic characteristics but is at least partly influenced by host-related factors.

TAC/SARIFA has recently emerged as a robust and easily assessable histomorphological biomarker in gastrointestinal malignancies. Several studies have demonstrated its prognostic relevance in colorectal and gastric cancer, and independent validation cohorts have confirmed its reproducibility and clinical significance (14, 18, 19, 21). The present study further supports these observations, as patients with at least one TAC/SARIFA-positive tumour showed significantly inferior overall survival. However, the primary objective of the current analysis was not to further validate the prognostic value of TAC/SARIFA, but rather to gain insight into its biological determinants.

Previous investigations suggest that TAC/SARIFA is not primarily driven by tumour genetics. Reitsam et al. showed the absence of specific genomic alterations associated with TAC/SARIFA positivity, arguing against a predominantly tumour-cell-intrinsic mechanism (14). In contrast, several studies have identified alterations of the immune microenvironment in TAC/SARIFA-positive tumours, including changes in natural killer cell populations and evidence of impaired anti-tumour immune responses (17–19). Together, these findings raise the possibility that TAC/SARIFA represents a host-conditioned tumour phenotype rather than a purely local histological phenomenon.

To investigate this hypothesis, we employed a within-patient design using synchronous and metachronous double and triple colorectal carcinomas. Such approaches are particularly informative when host-related determinants are suspected, because relevant patient-specific factors, including genetic background, systemic immune status, metabolic conditions, environmental exposures and lifestyle-related influences, are inherently matched within an individual (23) (24). At the same time, this design also has limitations, especially when temporal effects or biological interactions between tumour sites cannot be excluded (24). Nevertheless, analysing multiple independent tumours within one patient provides a suitable framework to test whether TAC/SARIFA status is associated with patient-related predisposition rather than only with tumour-intrinsic properties.

This question is particularly relevant in colorectal cancer, because synchronous carcinomas are uncommon and metachronous carcinomas are likewise relatively rare (25, 26). Accordingly, assembling a cohort of double and triple colorectal cancers required screening the archives of two large academic centres over a long time period. Although the proportion of synchronous and metachronous cases in our cohort was broadly in line with the published literature, the overall number of identified cases was lower than expected. This is most likely explained by the fact that case identification was based on full-text report queries rather than registry-based extraction. Therefore, some degree of selection bias cannot be completely excluded.

Interestingly, the postulated host-related pattern became apparent only after restricting the analysis to synchronous locally advanced tumours. In the overall cohort, TAC/SARIFA distribution did not differ from random expectation. This is likely explained by the inclusion of tumour constellations that are only partially informative for TAC/SARIFA biology, particularly early-stage pT1/pT2 lesions in which direct tumour-adipocyte contact rarely occurs, as well as metachronous tumours separated by time-dependent changes in the host milieu. Because TAC/SARIFA requires direct contact between tumour cells and adipocytes, its occurrence is intrinsically uncommon in early-stage carcinomas and therefore contributes little to the assessment of concordance within an individual patient. In contrast, synchronous pT3/pT4 carcinomas represent the biologically relevant setting in which TAC/SARIFA can occur and be meaningfully compared between tumours of the same patient. Within this subgroup, the observed concordance significantly exceeded the expected random distribution, supporting the existence of patient-specific factors promoting or preventing TAC/SARIFA formation.

These findings are also of interest in light of recent epidemiological observations. In the Netherlands Cohort Study, elevated body weight before tumour development was associated with a higher probability of TAC/SARIFA-positive colorectal cancer (27). By contrast, earlier retrospective studies that assessed obesity at the time of diagnosis did not identify such an association (15, 28). Taken together, these data suggest that systemic metabolic conditions preceding tumour development may contribute to the establishment of a tumour microenvironment permissive for direct tumour-adipocyte interaction. Obesity-related alterations in lipid metabolism, chronic low-grade inflammation and immune dysregulation are plausible candidate mechanisms linking host characteristics to TAC/SARIFA formation (29).

The absence of a comparable concordance pattern in metachronous tumours does not necessarily argue against this interpretation. Host-related determinants relevant to TAC/SARIFA formation are likely dynamic rather than static. Body weight, metabolic status, systemic inflammation, immune competence, microbiome composition and treatment-related effects may all change during the interval between two independent tumours. Such temporal variability may obscure patient-specific patterns and contribute to the random distribution observed in metachronous disease.

From a biological perspective, TAC/SARIFA may therefore be interpreted as a morphological surrogate of a specific tumour microenvironmental state characterised by permissive tumour-adipocyte interaction, altered immune surveillance and metabolic reprogramming. This interpretation is consistent with previous work showing associations between TAC/SARIFA and immune alterations as well as protease-related tissue remodelling (17, 19, 30). Future studies integrating spatially resolved molecular analyses and functional microenvironmental profiling will be needed to clarify the underlying cellular and molecular mechanisms.

Several limitations of the present study should be acknowledged. First, despite representing a comparatively large cohort of double and triple colorectal carcinomas, the number of synchronous locally advanced cases remained limited. Second, the retrospective design introduces the possibility of selection bias. Third, no functional analyses were performed, and mechanistic conclusions therefore remain speculative. Accordingly, the present findings should be regarded as hypothesis-generating and require validation in independent cohorts.

In conclusion, the present study provides evidence that TAC/SARIFA status is partially determined by host-related factors. The significant concordance observed among synchronous locally advanced colorectal carcinomas supports the concept that tumour-adipocyte interaction reflects a patient-specific biological predisposition rather than a purely tumour-intrinsic characteristic. These findings further strengthen the concept of TAC/SARIFA as a biologically meaningful biomarker and provide a rationale for future studies addressing the metabolic and immunological basis of tumour-adipocyte interaction.

## Data Availability

All data produced in the present study are available upon reasonable request to the authors

## 5 Acknowledgements

The authors are thankful to Barbara Köpf and Vera Popova of the Bavarian Cancer Registry (Landesamt für Gesundheit und Lebensmittelsicherheit) for providing the follow-up data. Furthermore, they are thankful to Kathrin Ferstl-Blahetek and her team for excellent technical work.

---

**Supplementary Table 1.**

|  | **All**<br>**n=276** | **SARIFApos**<br>**n=87** | **SARIFAneg**<br>**n=189** | **P-Value** |
| --- | --- | --- | --- | --- |
| Collect Würzburg | 75 | 24 | 51 | 0.917 |
| Collect Augsburg | 201 | 63 | 138 |  |
| Mean Age (± SD) | 70±12 | 70 ±12 | 70±12 | 0.406 |
| Sex (M : F) |  | 1.6:1 | 2.1:1 | 0.269 |
| Right Colon | 121 | 37 | 84 | 0.161 |
| Left Colon | 83 | 33 | 50 |  |
| Rectum | 58 | 12 | 46 |  |
| Total Colon/Rectum | 13 | 5 | 8 |  |
| Unknown Location | 1 | 0 | 1 |  |
| pT1 | 44 | 1 | 43 | **<0.001** |
| pT2 | 55 | 3 | 52 |  |
| pT3 | 140 | 62 | 78 |  |
| pT4 | 37 | 21 | 16 |  |
| pN0 | 160 | 33 | 127 | **<0.001** |
| pN+ | 109 | 54 | 55 |  |
| low grade | 213 | 62 | 151 | **0.015** |
| high grade | 57 | 26 | 31 |  |
| Tumor Budding 1 | 207 | 51 | 156 | **<0.001** |
| Tumor Budding 2/3 | 69 | 36 | 33 |  |
| MSS / pMMR | 165 | 60 | 105 | 0.071 |
| MSI / dMMR | 38 | 8 | 30 |  |
| Neo-Adjuvant Therapy | 21 | 3 | 18 |  |

SD = 1 standard deviation, M = male, F = female, MSS = microsatellite stable, MSI = microsatellite instable, pMMR = proficient mismatch repair, dMMR = deficient mismatch repair,

